# Digital Mental Health Support Reduces Intended Use of Clinical Services: An Observational Study of the Mindstep Platform

**DOI:** 10.1101/2025.06.23.25330119

**Authors:** Elisha Citron, Hamzah Selim, Aaron Lin

## Abstract

**Background:** The demand on mental health services is rapidly increasing, outpacing the capacity of traditional treatment systems like the NHS. Digital interventions offer a scalable means of addressing this gap, but real-world evidence of their impact on service utilisation is limited.

**Aims:** This study examines whether use of the Mindstep platform, a digital mental health tool offering assessments and video-based self-guided support, is associated with reduced intended use of clinical services and user-perceived usefulness.

**Methods:** A retrospective observational study was conducted using user survey data (N = 254) from individuals with at least 30 days of Mindstep exposure. Participants reported past-year healthcare use, rated the perceived usefulness of Mindstep, and estimated their future clinical service use both with and without Mindstep access. The primary outcome was changes in intended use of public and private clinical services; the secondary outcome was perceived usefulness.

**Results:** Mindstep use was associated with a 58.1% average reduction in intended clinical service use across all categories, with the largest reductions for private consultants and GPs. Users with low prior healthcare engagement reported the greatest reductions, while those with high prior usage rated Mindstep as most useful (mean usefulness score = 0.40 on a 0-1 scale).

**Conclusions:** Engagement with the Mindstep platform was associated with substantial reductions in intended clinical service use and moderate perceived usefulness, particularly among underserved and high-need user groups. While intention may not directly translate to behaviour, these findings suggest that digital mental health platforms may play an important role in alleviating demand on healthcare systems and improving access to early support.

## Introduction

Rising prevalence of common mental health conditions such as anxiety, depression, and attention-deficit/hyperactivity disorder (ADHD) has outpaced the capacity of traditional clinical services, contributing to record wait times across primary care, talking therapies, and emergency departments (NHS Digital, 2022). While evidence-based treatments exist, their provision remains constrained by systemic bottlenecks such as workforce shortages, limited commissioning capacity, and escalating demand, all of which have been further exacerbated by the lasting mental health consequences of the COVID-19 pandemic (McManus, et al. 2016; Pierce, et al. 2020).

Against this backdrop, digital mental health interventions have emerged as a potentially scalable means of extending support beyond the confines of the National Health Service (NHS; Hollis, et al. 2017; Firth, et al. 2017). However, robust evidence demonstrating their impact on healthcare utilisation in real-world UK populations remains limited. In particular, it is unclear whether such tools can meaningfully alter user behaviour or reduce reliance on strained services, especially outside of controlled trial environments.

*Mindstep* is a mobile web-based platform developed to bridge this gap between clinical need and access. Built around structured assessments and video-based psychoeducational interventions, the platform aims to serve both as a triage mechanism and as a source of ongoing, self-guided mental health support. Unlike symptom-specific apps, *Mindstep* uses dynamic branching logic and clinically informed design to tailor its guidance to users’ reported difficulties, including low mood, cognitive fog, burnout, and stress. Prior research has demonstrated the utility of *Mindstep’s* AI-driven assessments for early detection of cognitive impairment and for navigating users to appropriate levels of care, particularly in primary care settings (Alim-Marvasti, et al. 2022; Kuleindiren, et al. 2022).

In this study, we use a retrospective analysis design to examine *Mindstep’s* effectiveness by measuring reductions in intended use of clinical services and reported usefulness in addressing mental and brain health concerns across a real-world user base. Importantly, we assess intent across both public (e.g., NHS GPs and accidents and emergencies [A&E]) and private (e.g., private GPs and consultants) sectors, offering a rare lens on how digital tools interact with parallel care economies. This dual focus is particularly relevant in the UK, where rising demand has led many patients to seek faster – but often more costly – private alternatives (Care Quality Commission, 2024). By evaluating *Mindstep*’s influence across these pathways, we not only estimate its potential to alleviate pressure on the public health system, but also explore its role in reshaping access, affordability, and user autonomy within a hybrid healthcare landscape. We further stratify results by prior clinical engagement to understand how the platform may differentially affect high-frequency users versus individuals historically disengaged from care.

## Methods

### Participants

All users of the *Mindstep* app were eligible to complete the embedded user experience (UX) survey. However, only individuals with at least 30 days of *Mindstep* exposure in the last 12 months were included in the analysis. This criterion ensured that all responses reflected meaningful engagement with the platform across the same period of observation as used for service-use estimates. A total of 254 users submitted complete and valid responses and were retained for analysis.

Participants were stratified based on their historical clinical service engagement:

- Primary care visits: categorised as 0 visits, 1–2 visits, 3–5 visits, 6–9 visits, and ≥10 visits.
- Previous emergency (A&E) service utilisation: Yes vs. No.
- Previous use of talking therapy: categorised as 0 visits, 1–2 visits, 3–5 visits, 6–9 visits, and ≥10 visits.

### Design

To examine whether *Mindstep* use was associated with changes in users’ intended reliance on clinical services, this study employed a retrospective, counterfactual survey design. The analysis was structured to assess service use over a 24-month window – the previous 12 months, during which participants were introduced to Mindstep, and the prospective 12 months.

Previous service use was assessed based on participants’ reported frequency of visits to three service types: PCDs, emergency departments (A&E), and talking therapists. In the UK context, PCDs typically refer to general practitioners (GPs), who may be accessed through the public NHS or privately. A&E refers to NHS emergency departments providing urgent care for acute or life-threatening conditions. Talking therapists are mental health professionals who offer psychological support through verbal interventions and may operate within either NHS or private settings.

For the purposes of outcome analysis, clinical services were grouped into five categories: (1) NHS General Practitioners (public PCDs), (2) Private General Practitioners (non-NHS PCDs), (3) NHS A&E services, (4) Private consultants (non-NHS specialist doctors such as psychiatrists), and (5) the NHS 111 service – a free, 24/7 phone and online service designed to help individuals access appropriate care for urgent but non-emergency health concerns. NHS services (GPs, A&E, and 111) were classified as public, while private GPs, consultants, and non-NHS therapy providers were categorised as private. This classification reflects typical distinctions within the UK healthcare system but may differ in international contexts.

### Materials

The survey (Appendix 1) consisted of six structured questions designed to capture:

- Historical clinical service use over the past 12 months (primary care doctors [PCDs], talking therapists, and emergency services).
- Perceived usefulness of *Mindstep*, measured on a five-point ordinal scale ranging from “not useful at all” to “extremely useful.”
- Use of clinical services in the next 12 months if *Mindstep* had not been available (counterfactual scenario).
- Use of clinical services over the next 12 months, considering continued access to *Mindstep* (measured on an ordinal likelihood scale).

Ordinal response categories were converted into numeric values to facilitate statistical analyses. In the absence of corresponding behavioural data, this was done based on *a priori* semantic meaning (Appendix 2). Future intended use and perceived usefulness responses were quantified on a continuous scale from 0 – 1. Past service-use frequencies were categorised using midpoint values for interval-based responses (e.g., “3–5 times” assigned a value of 4).

### Procedure

The primary outcome measure was the reduction in intended use of clinical services, defined as the proportional change between users’ counterfactual estimates (without Mindstep) and their intended use given access to Mindstep. By comparing estimates of future intended use and a counterfactual baseline, we were able to assess proportional changes in service use attributable to *Mindstep* in a real-world, post-deployment setting. This was calculated as:

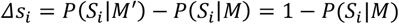

where:

- Δ*s*_*i*_ denotes the change in intended future use of clinical service *S*_*i*_ (e.g., GP, therapist, emergency services)
- *M* represents engagement with the *Mindstep* app
- *M*′ represents the counterfactual scenario without *Mindstep*
- *P*(*S*_*i*_|*M*) is the reported probability (or self-rated likelihood) of using service *S*_*i*_ in the future given access to *Mindstep*.

Under the assumption that participants would have definitely used a given service in the absence of Mindstep (i.e., *P*(*S_i_*|*M*′) = 1), this formulation simplifies to a direct measure of proportional reduction attributable to Mindstep exposure. Mean reductions were computed for each clinical service category as well as pooled across all services. While self-reported intention is not a definitive proxy for behaviour, it offers a pragmatic and scalable early signal that can guide experimental validation where direct outcome tracking may be infeasible.

The secondary outcome measure was the perceived usefulness of *Mindstep*, scored numerically from ordinal responses. This item captures subjective evaluations of the app’s impact across a range of user experiences, providing insight into users’ perceived value of the intervention, which may also be relevant to future service use.

Statistical analyses were conducted in Python (v3.11.2) using the `pandas, numpy, stats, matplotlib.pyplot` and `scipy.stats` libraries. Error bars were calculated using the standard error of the mean (SEM).

Inferential statistics included:

- Paired-sample t-tests (for differences in intended use)
- One-way ANOVAs (for subgroup comparisons across prior usage bins)
- Pearson and Spearman correlations (for linear associations)

All collected data were anonymised and gathered through voluntary in-app surveys as part of ongoing product evaluation. Ethical approval for studies involving Mindstep data was originally obtained from the NHS Health Research Authority (West Midlands Solihull Research Ethics Committee) on 16/09/21. This data was collected as post-market research for the purpose of product refinement.

## Results

### 1. Sample Characteristics

Of the total respondents to the *Mindstep* user experience survey, N = 254 users provided complete data for analysis. Participants represented a wide range of prior clinical engagement:

- 52.0% reported visiting a PCD for mental or brain health in the past 12 months.
- 48.0% had accessed talking therapy.
- 7.48% had presented to A&E for mental health concerns.

Demographic information was not collected.

### 2. Overall Changes in Intended Service Use and Perceived Usefulness

Following *Mindstep* use, users reported a substantial drop in their intention to access clinical services (Figure 1). Across all service categories, users reported statistically significant reductions in intended future use (*p* < .0001), with a mean reduction of 58.1%. The largest reductions were in intended visits to private consultants (65.5%; *t* = 13.79) and private GPs (60.0%; *t* = 12.05). NHS services also showed a significant reduction with NHS GPs dropping by 57.1% (*t* = 24.13) and A&E visits by 52.5% (*t* = 8.05).

**Figure 1.**
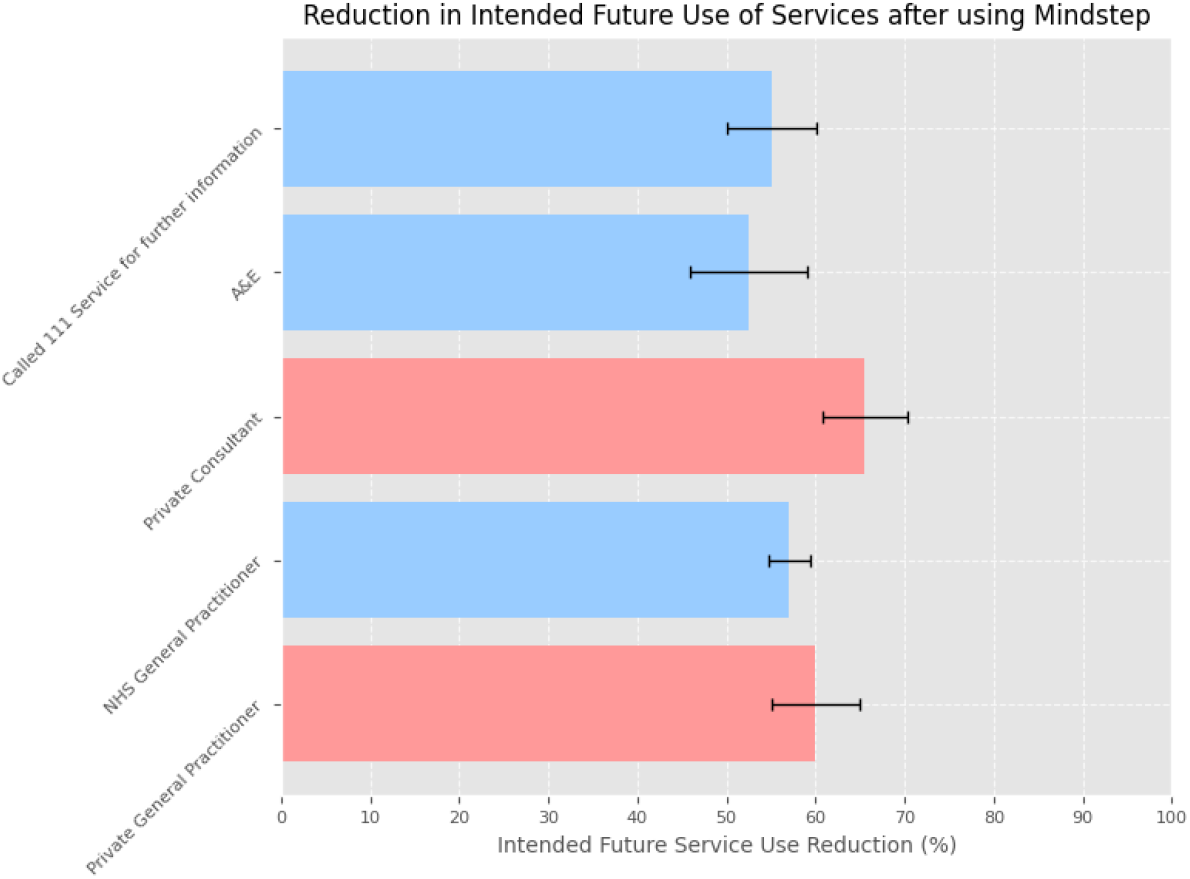
Bar chart showing mean % reduction in intended clinical service use. Private services are shown in red while NHS services are shown in blue. Y-axis: Clinical service type. X-axis: % reduction in intended use.

User ratings indicated generally favourable perceptions of *Mindstep’s* utility, with 50.4% of users rating the app as at least “moderately useful” (Figure 2). On a 0–1 scale (mapped from five ordinal categories), the mean usefulness score was 0.40 (± 0.018).

**Figure 2.**
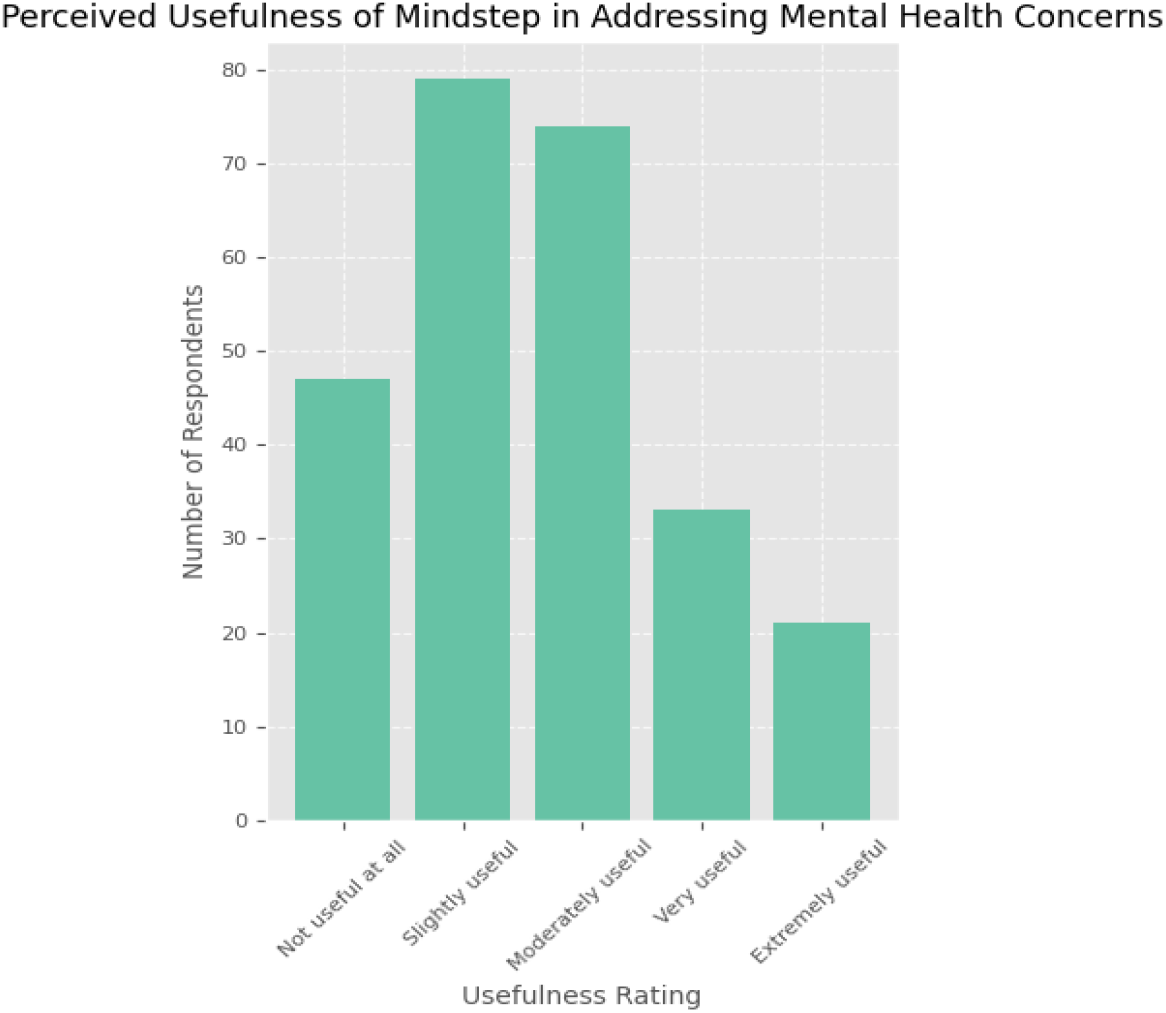
Bar chart of perceived usefulness scores. Y-axis: Number of users. X-axis: Usefulness categories.

### 3. Impact of Prior Primary Care Use on Service Reduction and Usefulness

Service reduction effects varied significantly by baseline usage, with users who rarely saw their PCD showing the greatest reductions (Figure 3; ρ = –0.90, p = 0.037). For example, users with no PCD visits in the past year reported a 66.3% average reduction across all services, while those with ≥10 visits reported only 37.8%. A one-way ANOVA comparing reduction percentages across prior PCD visit frequency bins (0, 1–2, 3–5, 6–9, ≥10 visits) revealed a significant effect of frequency, *F*(4, 249) = 7.13, *p* < 0.001.

**Figure 3.**
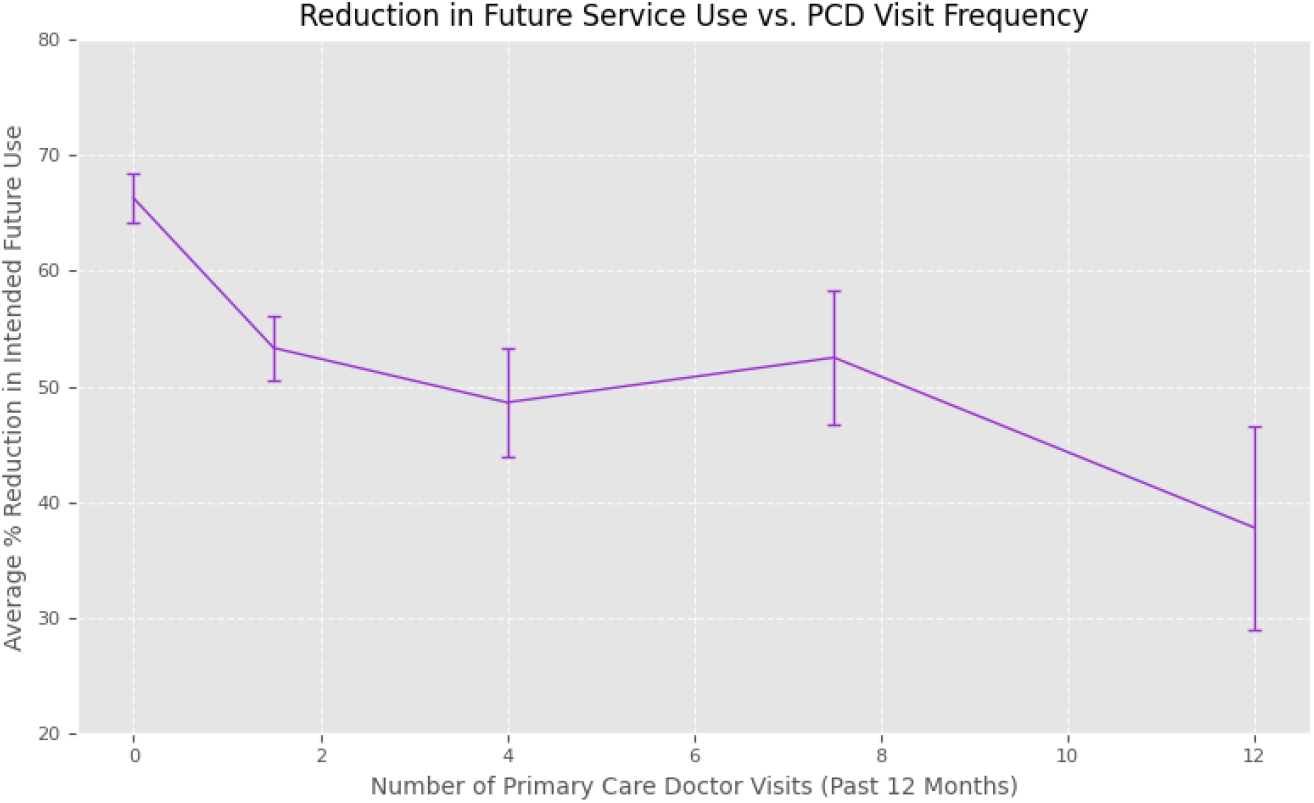
Line graph showing reduction in intended service use vs. prior PCD visits. X-axis: Number of prior PCD visits. Y-axis: % reduction in intended service use.

In contrast, users with higher prior PCD use reported higher perceived usefulness, with the mean usefulness score being 0.25 points higher among frequent users (≥10 visits) than among non-users. (Figure 4; *ρ* = 1.00, *p* < 0.001). A one-way ANOVA confirmed a significant effect of visit frequency on perceived usefulness, *F*(4, 249) = 3.02, *p* = 0.019.

**Figure 4.**
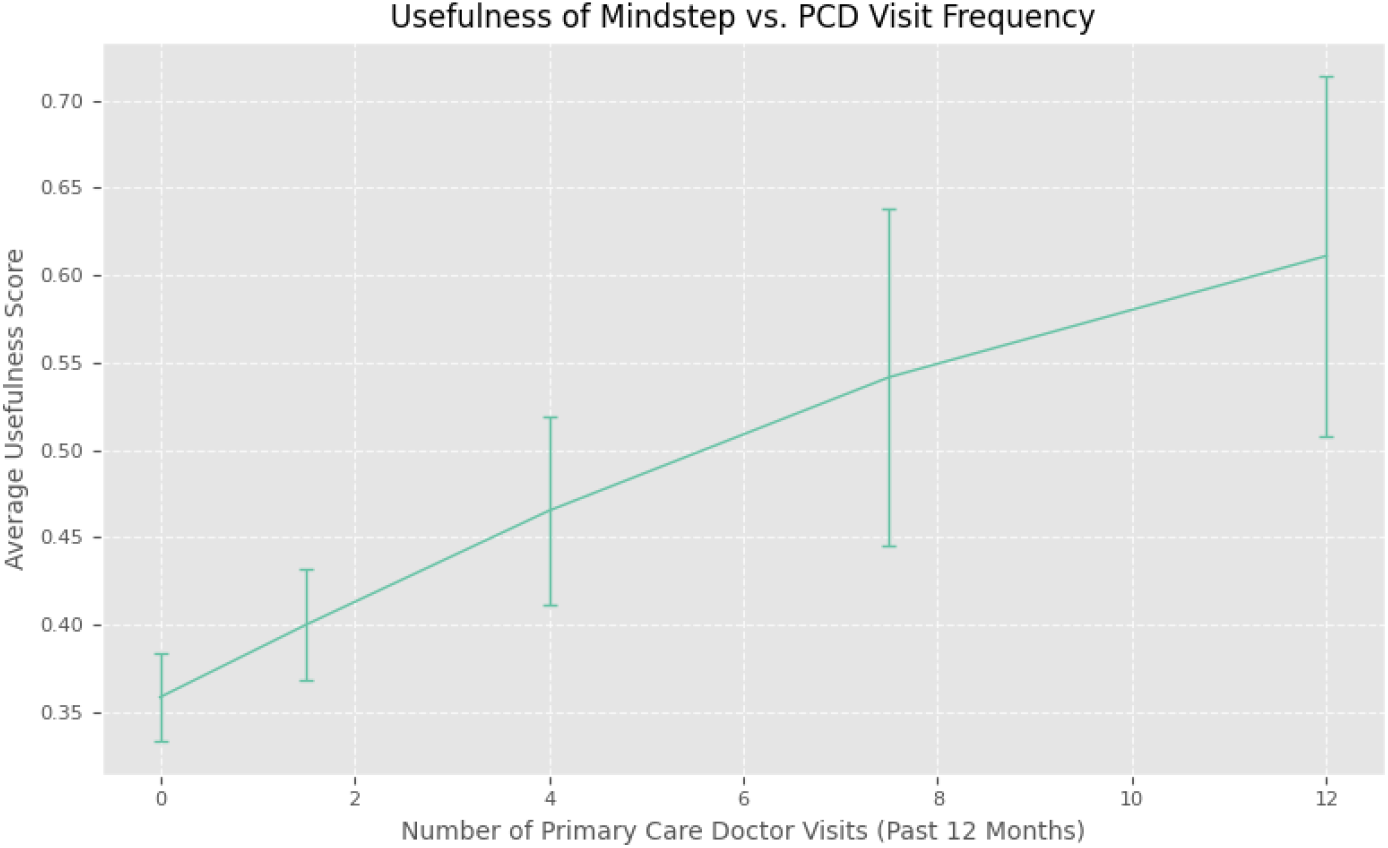
Line graph showing usefulness score vs. prior PCD frequency. X-axis: Number of prior PCD visits. Y-axis: Mean usefulness score (0–1 scale).

### 4. Influence of Prior Emergency Service Use on Future Intentions

There was no significant difference in general future service use reduction between participants who had visited A&E for their mental health over the past 12 months and those who had not (*t* (22.02) = 0.9; *p* = 0.37). However, isolating distinct services revealed that future intended A&E use was particularly impacted by *Mindstep* use in users who had visited A&E one or more times in the past 12 months (Figure 5; *t* (7.23) = -2.08; *p* = 0.07). Among users with past A&E attendance, intended future A&E use dropped by 72.5%, markedly higher than the 50.0% reduction among those without prior A&E use.

**Figure 5.**
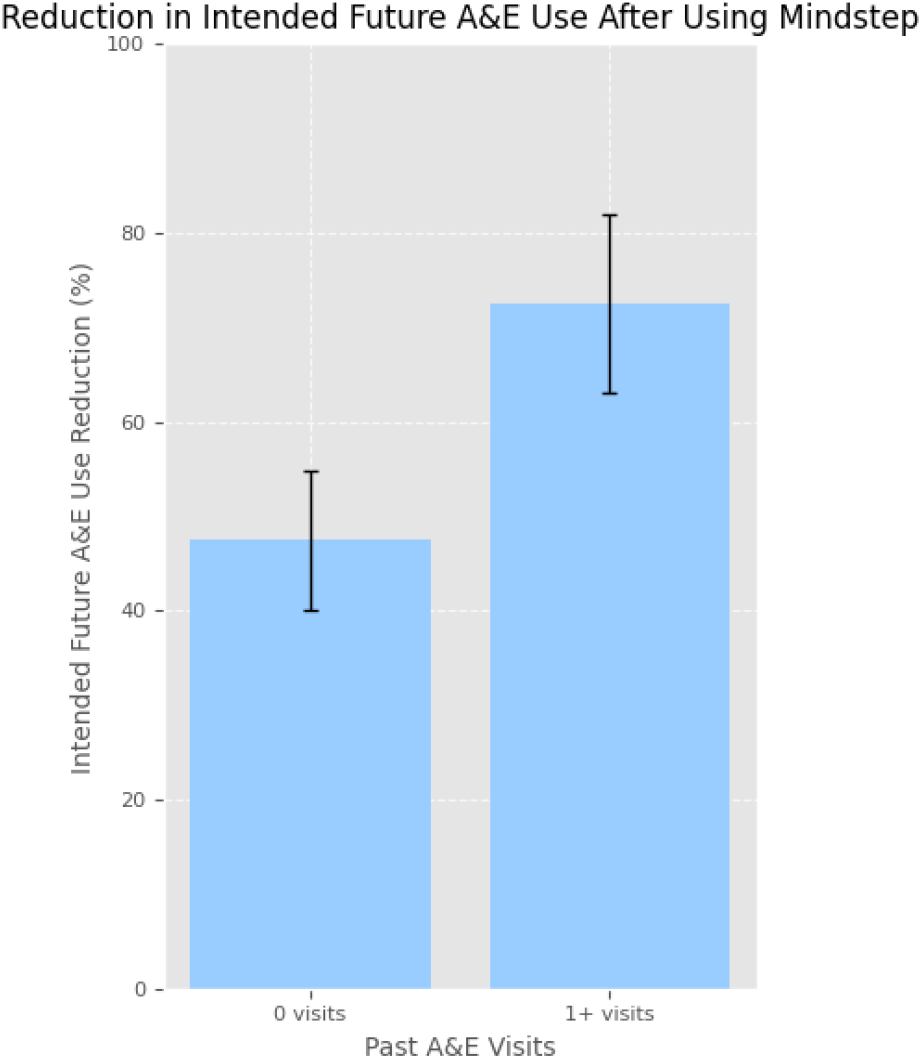
Bar chart comparing % reduction in A&E use between users with vs. without prior A&E attendance. X-axis: Group (Past A&E user, No A&E use). Y-axis: Mean % reduction.

However, there was no significant difference in perceived usefulness of *Mindstep* between users who had and had not visited A&E during the past 12 months (*t* (3.77) = 0.56; *p* = 0.61).

### 5. Relationship Between Past Therapy Use and Reported Outcomes

User’s history of talking therapy visits showed a significant negative association with the reduction in future intended clinical service use (Figure 6; ρ = –0.70, p = 0.019). The shape of this plot closely mirrors that of Figure 3: an initial steep decline, followed by a plateau, a slight increase, and a final drop. This parallel trend warrants further investigation, though it may largely reflect overlap between user groups, given that many respondents had seen both a primary care doctor and a talking therapist in the past 12 months. However, the overall magnitude of this relationship was smaller than that observed for PCD use, suggesting that prior engagement with talking therapy may exert a weaker influence on how users respond to Mindstep in terms of intended service reduction.

**Figure 6.**
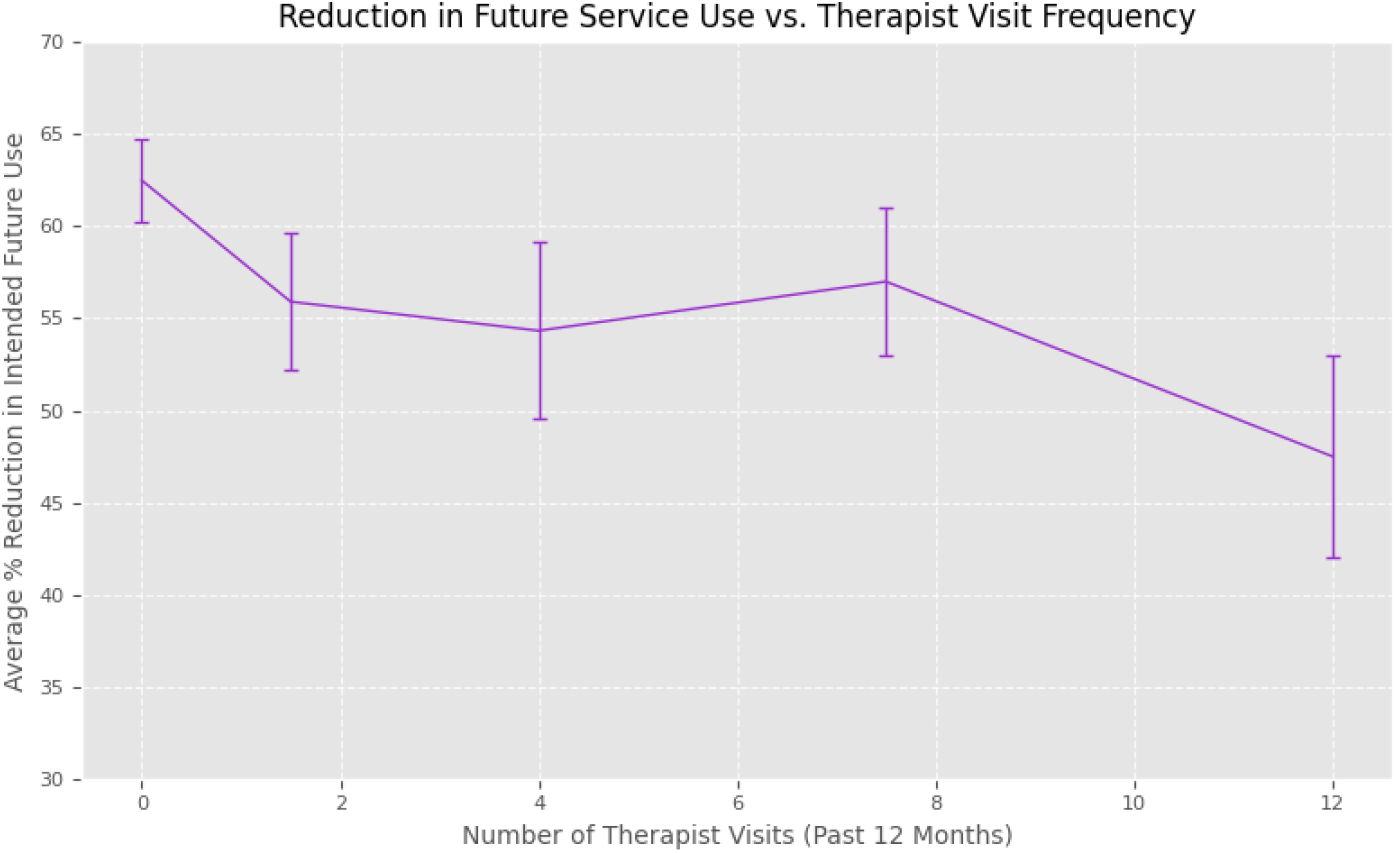
Line graph showing reduction in intended service use vs. prior talking therapist visits. X-axis: Prior visit frequency. Y-axis: % reduction in intended service use.

Finally, similar to A&E-based groupings, prior talking therapist visit frequency had no significant effect on perceived usefulness of *Mindstep* (*ρ* = 0.10, *p* = 0.873).

## Discussion

This study provides early, real-world evidence that digital mental health interventions such as *Mindstep* may substantially reduce users’ intended reliance on clinical services across primary, secondary, and emergency care. This is reflected by a 58.1% average reduction in anticipated service use following app engagement, with particularly pronounced declines in intended visits to private consultants and GPs. These findings validate broader efforts to decentralise mental health care and support a stepped care model, in which digital tools serve as a scalable first line of support.

A key insight emerging from the data is the differential impact of *Mindstep* based on prior clinical engagement. Users with no recent history of primary care or talking therapy visits reported the greatest reductions in intended service use, suggesting that the platform may be particularly valuable for hard-to-reach populations who are underserved by traditional systems. This supports the hypothesis that digital tools can extend the reach of mental health care to those reluctant or unable to seek in-person help.

In addition, this finding may support the complementary hypothesis that platforms like Mindstep enable users to access support earlier in the course of their mental health difficulties, before symptoms escalate to the point of requiring clinical intervention. By lowering barriers to entry, such tools may encourage proactive engagement, helping users self-manage or address concerns at an earlier stage, potentially averting future deterioration and reducing demand on overstretched services. This preventative dimension, while harder to measure directly in an intention-based study, may represent a key mechanism through which digital interventions contribute to population-level mental health outcomes.

The observed 72.5% reduction in intended A&E use among those with prior emergency visits is particularly notable, given the resource-intensiveness and often avoidable nature of many psychiatric presentations to urgent care. While intention does not guarantee behaviour, this finding offers a promising early signal that *Mindstep* may deflect high-risk users from acute care pathways by providing timely, low-friction support to prevent future urgent treatment.

This study also revealed several interesting trends regarding users’ self-reported usefulness of the *Mindstep* platform.

In contrast to findings of increased service use reductions, participants with high prior healthcare usage reported greater perceived usefulness of *Mindstep*. This discrepancy may imply a complementary rather than substitutive role but warrants further exploration. One possible explanation is that it may reflect the role of digital interventions in augmenting rather than replacing formal care for individuals with complex or chronic mental health conditions. It is also notable that no such relationship between prior use and usefulness was found for A&E or talking therapy. This may suggest that *Mindstep’s* perceived value is most salient among individuals engaged in generalist or front-line care, where digital tools can supplement guidance, monitoring, or triage. In contrast, users engaged in specialist or crisis-oriented services - such as psychotherapy or emergency care - may have needs or expectations that digital self-help tools alone cannot fulfil.

A noteworthy dimension of this study is its inclusion of both public (e.g., NHS GP, A&E) and private (e.g., private GP, consultants) healthcare services in its analysis of intended use. The finding that *Mindstep* users reported the largest reductions in planned engagement with private providers (67.5% for consultants and 64.8% for private GPs) compared to NHS services (55.6% for GPs and 54.7% for A&E) suggests that digital interventions may exert a stronger substitutive effect within the private sector. This likely reflects not only the cost-sensitivity of self-paying users but also the potential for digital tools to reduce discretionary spending on mental health consultations where self-management is perceived as sufficient. At the same time, the meaningful reduction in NHS-facing demand indicates that *Mindstep* may also serve a system-level role in alleviating pressure on public services. The dual utility across care economies underscores the potential of digital platforms to operate flexibly within mixed healthcare systems, offering both economic and capacity benefits depending on the user’s context and care-seeking pathway.

Nonetheless, several limitations must temper the interpretation of these findings. The study’s reliance on intention data, rather than observed behaviour, limits causal inference and introduces potential biases such as social desirability or recall distortion. Additionally, the use of intuitive numerical mappings to translate ordinal responses, though practical for exploratory analysis, may obscure meaningful subtleties in user perception and should be refined through psychometric validation. The absence of demographic data further restricts the ability to generalise these findings or explore equity implications which are both critical in the context of digital health adoption.

Future research should prioritise linkage of app data to real-world healthcare utilisation metrics to confirm behavioural impact. Experimental designs such as randomised controlled trials (RCTs) or quasi-experimental studies using matched controls would provide more definitive evidence of causal effects. Furthermore, stratified analyses across demographic groups and clinical profiles are essential to identify which users benefit most - and in what ways - from tools like *Mindstep*.

## Conclusion

Taken together, these findings support the hypothesis that thoughtfully designed digital mental health platforms can reduce intended clinical service demand, particularly among first-time or infrequent healthcare users. While not a replacement for formal care, *Mindstep* and similar tools may serve as a vital layer in a modern, adaptive mental health ecosystem - one that meets individuals earlier, and more often, in their journey toward wellbeing.

## Funding

Funding for this paper was provided by Mindset Technologies Ltd.

## Conflict of Interest

All authors are affiliated with Mindset Technologies Ltd., the developer of the Mindstep platform evaluated in this study.

## Data Availability

All data produced in the present study are available upon reasonable request to the authors.

## Appendices

Appendix 1: Survey Questions

1. In the past 12 months, how often have you seen your Primary Care doctor (Private or NHS GP) for your mental health?
2. In the past 12 months, how often have you seen a talking therapist (including telephone or online) for your mental health?
3. In the past 12 months, how often have you attended emergency services (specifically, A&E/Urgent care) for your mental health?
4. To what extent has Mindstep been helpful in addressing your mental or brain health concerns?
5. If Mindstep was not available, where would you have gone for help with your mental health concerns? Please select all that apply.
6. Now that you have used Mindstep, how often do you think you will use the same service (i.e. service you specified above) in the coming 12 months?

Appendix 2: Numerical Mappings

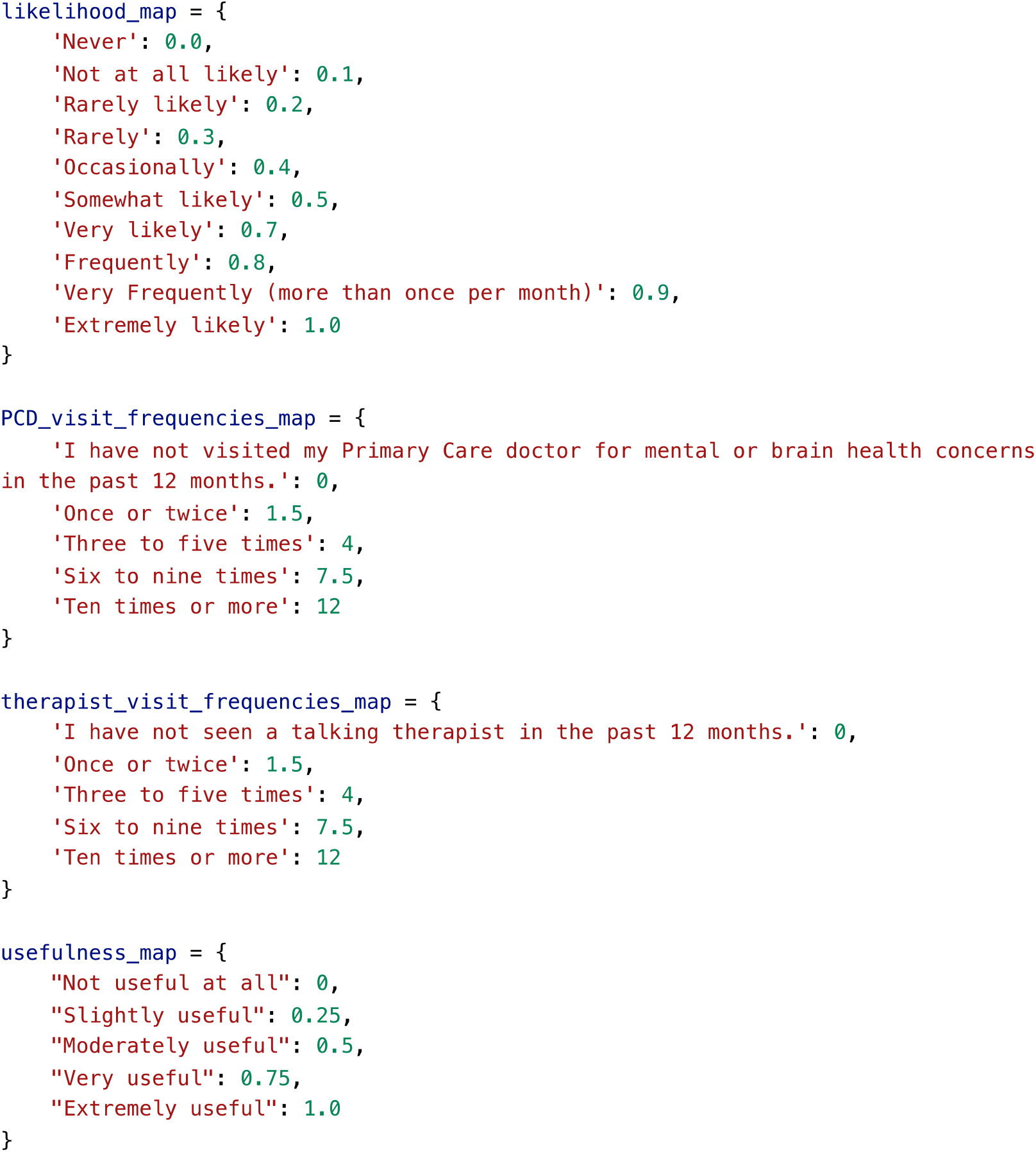

## Notes

### Author Declarations

Ethical approval for studies involving Mindstep data was originally obtained from the NHS Health Research Authority (West Midlands Solihull Research Ethics Committee) on 16/09/21. This data was collected as post-market research for the purpose of product refinement.

